# Percentile Scores for the Revised University of Pennsylvania Smell Identification Test for 16,972 Individuals 60 years of Age and Older

**DOI:** 10.1101/2024.10.01.24314727

**Authors:** Kerri A. Pierz, Whitley Aamodt, Caroline Gochanour, Ryan Kurth, Michael C. Brumm, Christopher S. Coffey, Laura E. Heathers, Michelle Totten, Richard L. Doty, Kenneth Marek, Andrew Siderowf, the Parkinson’s Progression Markers Initiative

## Abstract

**Background and Objectives:** The University of Pennsylvania Smell Identification Test (UPSIT) is a widely employed objective measure of smell function. A revised version of this test, termed the UPSIT-R, was developed in 2020 to update certain odorants and response options. The goal of this study was to establish normative age- and sex-specific data for the UPSIT-R in a large adult cohort and to compare the resultant percentiles to those of the original UPSIT. A second study was performed to compare the relative performance of these two tests in a small cohort of persons with and without Parkinson’s disease (PD).

**Methods:** UPSIT-R percentiles were derived separately by age (60-64, 65-69, 70-74, 75-59, and ≥80 years) and sex (male, female) categories in 16,972 volunteers who were predominantly white and non-Hispanic. Percentiles were derived statistically using an empirical distribution averaging procedure. A Bland-Altman plot was employed to visualize the level of agreement between percentiles for the UPSIT-R and UPSIT. For the second study, non-parametric statistics were employed to compare the results of those with and without PD.

**Results:** UPSIT-R performance declined with increasing age; deficits were more pronounced in men than in women. The magnitude of the difference between the original and revised test percentile scores differed by age and sex; women had up to a 3-point improvement with the UPSIT-R whereas the oldest male age had up to a 3-point decline in percentile scores. Olfactory deficits in PD were confirmed on the UPSIT-R.

**Discussion:** This study provides normative data clinically useful for assessing the relative degree of dysfunction in persons 60 years of age and older using the UPSIT-R. Like the traditional version of the UPSIT, this version demonstrates that smell function declines with age and is generally better in women than in men, providing a solid clinical basis for physicians to accurately determine the degree of smell dysfunction in their patients.

**Trial Registration Information:** ClinicalTrials.gov NCT05065060

## Introduction

Olfactory dysfunction has been recognized as a marker of neurological disease, specifically with evidence from individuals with Parkinson disease (PD), Alzheimer disease, and myasthenia gravis.^1, 2, 3, 4, 5^

Historically, much research designed to understand the prognostic value of olfactory function for neurodegenerative diseases has utilized The University of Pennsylvania Smell Identification Test (UPSIT), a 40-item test that was originally developed in 1984.^6^

Previous studies have described the use of threshold values based on UPSIT score to assign olfactory diagnoses ranging from normosmia (UPSIT ≥34 in males; ≥35 in females) to total anosmia (≤18 for males and females).^7, 8^ The limitations of a classification approach based on absolute UPSIT scores have been previously discussed.^9^ We recently published updated age- and sex-specific UPSIT percentiles for adults 50 years of age and older based on a large combined sample from the Parkinson Associated Risk Syndrome (PARS) and Parkinson’s Progression Markers Initiative (PPMI) cohorts.^9^ While those percentiles remain valuable for interpretation of data from studies using the original form of the UPSIT,^6^ the instrument was revised in 2020 to modify some of the odorants and response distractors. Thus, as use of the revised form of the UPSIT becomes more widespread, it will be useful to understand the performance differences between the two versions of the test and to have age- and sex-specific percentile values for the revised UPSIT available that can inform data interpretation.

The research presented here was designed to explore differences between the original UPSIT from 1984 and the revised version (UPSIT-R) released in 2020. It is of importance to understand how the UPSIT-R compares in performance to the original UPSIT so that existing and future datasets can be appropriately interpreted and compared.

## Methods

### Bridging Study Participants and Design

Participants included healthy controls (HC) and people diagnosed with PD who completed both versions of the UPSIT at either Indiana University (IU) or the University of Pennsylvania (Penn). The bridging study used a within-subjects design to compare performance on the original and revised versions of the UPSIT.

At Penn, PD and HC participants between 50-89 years of age were recruited during routine clinical or research visits at the Parkinson’s Disease and Movement Disorders Center. PD participants had a known diagnosis of idiopathic PD, while control participants had no known neurodegenerative disease. Exclusion criteria were as follows: atypical parkinsonian syndrome, history of prior head trauma or traumatic brain injury, underlying sinus or nasal dysfunction, current upper respiratory tract infection, use of inhaled tobacco products within the preceding 6 months, or an inability to give consent as judged by the treating physician.

At IU, HC participants were recruited from a registry of individuals who had previously agreed to future contact for new PD research. Eligible subjects were ≥60 years of age, able to provide written informed consent, and able and willing to comply with online study procedures. Interested participants were provided a link to review and sign the informed consent online and complete the screening process. Signature of the informed consent was delayed until all questions by the participant were addressed by study staff.

Participants from both sites completed a standardized questionnaire to collect the following demographic and clinical information: age, sex, race/ethnicity, history of sinus or nasal dysfunction, prior tobacco use, and subjective hyposmia/anosmia and dysgeusia/ageusia. All participants were requested to complete both the original and revised versions of the UPSIT. Penn participants completed one version of the UPSIT in the office and the second version at home. IU participants completed both versions of the UPSIT at home. Participants were provided with stamped, self-addressed envelopes and instructed to return tests by mail within two weeks of their initial test date. At Penn, the order of UPSIT administration was determined by a random number generator, so that half of participants completed the original version in the office and the revised version at home and vice versa. At IU, all participants were administered the original UPSIT first, followed by the revised UPSIT. UPSIT raw scores were calculated for both test versions and converted to age/sex-adjusted percentile scores based on normative data derived from the original UPSIT. Each participant was assigned an ID number, and de-identified scores were stored in a password-protected database.

### Cohort Study Participants and Design

Two newly enrolled populations from PPMI Online and Smell Test (ST) Direct were utilized to generate new age- and sex-specific percentile scores for the revised version of the UPSIT. Raw scores and percentiles on the original UPSIT from the combined PARS and PPMI cohort previously reported^9^ were compared to those for the revised UPSIT collected from PPMI Online and ST Direct cohorts.

#### 1. PPMI Online

PPMI is a longitudinal, observational, multi-center natural history study of PD. PPMI Online refers to a large cohort of individuals recruited online, from which subgroups are further selected based on qualifying criteria to participate in various investigations of the progression of clinical features, imaging outcomes, or biologic and genetic markers possibly related to PD. For more information, see [study link](https://www.ppmi-info.org/study-design). In the study reported here, only data from participants in PPMI Online aged 60 years or older who did not have a PD diagnosis were included. Recruitment strategies included invitation to participate through other ongoing PPMI studies, Fox Insight, independently managed PD studies, digital and social marketing recruitment efforts to the PD community, and events/activities conducted by The Michael J. Fox Foundation or other representatives of PPMI. Participants completed a screening questionnaire and consented online using Evidation Health’s eConsent process prior to performing other study activities.

Participants completed online questionnaires to collect demographics and vital statistics, and medical and family history. An UPSIT (revised version) was provided in person or mailed to eligible participants to complete. Participants were instructed to submit test responses via a web-based portal. Enrollment into PPMI Online is ongoing with a long-term goal of recruiting 500,000 participants globally. PPMI Online is registered on clinicaltrials.gov as NCT05065060.^10^

#### 2. ST Direct

ST Direct seeks to identify individuals in the general population with an impaired sense of smell. This protocol engaged individuals from the general population aged 60 and older without a PD diagnosis living in the United States, Canada, England and the Netherlands. Participants were led to a web portal via QR code or URL where they were screened for eligibility. Contact information was collected, and consent was completed online. An UPSIT (revised version) was mailed to eligible participants to complete. Participants were instructed to submit test responses via a web-based portal.

Paid media and advertisements, collaborations with The Michael J. Fox Foundation and other organizations (including Quest Diagnostics, 23 and Me, Smell and Taste Association of North America), and active senior living outreach were some of the more successful recruitment efforts utilized as part of a multi-faceted approach. The study utilized a centralized, remote screening approach that de-emphasized direct site team outreach and made it easier for individual and community organization partners to refer participants. Enrollment into ST Direct is ongoing with a long-term goal of recruiting 500,000 participants.

### Standard Protocol Approvals, Registrations, and Patient Consents

All study participants provided written informed consent to participate in the associated studies. Bridging Study IRB approvals provided by IU IRB (protocols #1906568726 and 10630) and Penn IRB (protocol #843055); PPMI Online (protocol #20211908), ST Direct (protocol #12886). All research was conducted in accordance with the Declaration of Helsinki and the Good Clinical Practice guidelines after approval of the local ethics committees of the participating sites.

### Statistical Analysis

For the bridging study, descriptive statistics were calculated for the overall sample and by study group (HC vs. PD). Wilcoxon signed-rank tests were used to assess within-individual differences in UPSIT performance by version; Wilcoxon rank-sum tests were used to compare differences by study group and block order. To compare the revised and original UPSIT, the average UPSIT raw score and the difference in UPSIT percentiles were plotted using the “BlandAltmanLeh” package in R.^11, 12^

Within the cohort study, descriptive statistics were calculated separately by sex and by population (PPMI Online, ST Direct, and combined). UPSIT percentiles were derived separately by sex and age category (60-64, 65-69, 70-74, 75-59, and ≥80 years) using the default method (based on an empirical distribution function with averaging) in the UNIVARIATE procedure in SAS v9.4 (SAS Institute Inc., Cary, NC; sas.com; RRID:SCR_008567). Percentiles corresponding to each UPSIT raw score were rounded to the nearest integer. If a raw score corresponded to multiple percentile values, the median percentile was used. If a raw score fell between two percentiles, the upper and lower bordering percentiles were averaged and rounded up to the nearest integer. All scores that corresponded to a percentile less than 1 were rounded to the 1^st^ percentile.

### Data Availability

This analysis used data openly available from PPMI; data used in the preparation of this manuscript were obtained on July 3, 2023 from the Parkinson’s Progression Markers Initiative (PPMI) database (ppmi-info.org/access-data-specimens/download-data), RRID: SCR 006431. For up-to-date information on the study, visit www.ppmi-info.org. This analysis was conducted by the PPMI Statistics Core and used actual dates of activity for participants, a restricted data element not available to public users of PPMI data. Statistical analysis codes used to perform the analyses in this article are shared on Zenodo [10.5281/zenodo.11644053].

## Results

### Bridging Study: Within-subject comparison of performance between original and revised UPSIT Characteristics of the study population

The bridging study cohort included n=89 HC and n=48 PD participants recruited from IU and Penn. Demographics of this population are shown in Table 1. All PD patients were recruited from Penn and most (89%) of the HC participants were recruited from IU. The median age was 67.0 and 69.5 years in the HC and PD groups, respectively.

**Table 1.** Demographics of bridging study participants.

| Variable | Subgroup |  |  |
| --- | --- | --- | --- |
|  | Combined<br>(N = 137) | HC<br>(N = 89) | PD<br>(N = 48) |
| **Site, n (%)** |  |  |  |
| IU | 79 (58%) | 79 (89%) | 0 |
| Penn | 58 (42%) | 10 (11%) | 48 (100%) |
| **Block Order, n (%)** |  |  |  |
| Original before Revised | 108 (79%) | 84 (94%) | 24 (50%) |
| Revised before Original | 29 (21%) | 5 (6%) | 24 (50%) |
| **Sex, n (%)** |  |  |  |
| Female | 75 (55%) | 53 (60%) | 22 (46%) |
| Male | 62 (45%) | 36 (40%) | 26 (54%) |
| **Age at first UPSIT, mean (SD)** | 67.5 (6.2) | 67.2 (5.0) | 67.9 (8.1) |
| Median (Min, Max) | 67.0 (50, 86) | 67.0 (60, 86) | 69.5 (50, 80) |
| **Race (White), n (%) \*** | 122 (92%) | 83 (97%) | 39 (83%) |
| Missing | 4 | 3 | 1 |
| **Ethnicity (Hispanic or Latino), n (%)** | 1 (1%) | 1 (1%) | 0 |
| Missing | 4 | 4 | 0 |

\*Across the control and PD subgroups, 1 individual self-reported as Asian, 8 as Black or African American, and 2 as multiracial.

### Impaired olfaction in PD subjects compared to HC participants confirmed

As predicted, olfactory function was lower in the PD compared to the HC participants (Table 2). The median (IQR) UPSIT raw score for the original test was 17 (14-21.5) among PD and 34 (31-36) among HC participants. Similarly, the median (IQR) UPSIT score for the revised test was 19 (15-27) among PD and 36 (33-37) among HC participants.

**Table 2.** Comparison of average raw scores and percentiles between original and revised versions of the UPSIT.

|  | **HC**<br>(N = 89) | **PD**<br>(N = 48) |
| --- | --- | --- |
| **Raw scores** |  |  |
| Original UPSIT, mean (SD) | 32.7 (5.4) | 18.6 (7.3) |
| Median (IQR) | 34 (31-36) | 17 (14-21.5) |
| Revised UPSIT, mean (SD) | 34.4 (5.2) | 21.1 (8.5) |
| Median (IQR) | 36 (33-37) | 19 (15-27) |
| Difference (Revised – Original), mean (SD) | 1.7 (2.6) | 2.5 (4.5) |
| Median (IQR) | 2 (0-3) | 2 (1-5) |
| **Percentiles** |  |  |
| Original UPSIT, mean (SD) | 56.4 (28.0) | 10.2 (15.7) |
| Median (IQR) | 55.5 (39-80.5) | 4.5 (3-9) |
| Revised UPSIT, mean (SD) | 71.1 (28.4) | 18.1 (27.8) |
| Median (IQR) | 81 (50-95.5) | 6 (4-13) |
| Difference (Revised – Original), mean (SD) | 14.7 (19.9) | 7.9 (20.2) |
| Median (IQR) | 13.5 (1.5-27) | 2 (0-6) |

### Within-subject comparison of the original and revised UPSIT raw scores and percentiles

Higher scores on the revised UPSIT were observed in 102 (74%) of participants; 23 (17%) had lower scores on the revised UPSIT; 12 (9%) scored the same on both versions. Wilcoxon signed rank tests indicated significantly higher raw scores on the revised UPSIT than the original UPSIT, both among HCs (*P* <.0001) and those with PD (*P* <.0001); median (IQR) differences were 2 (0-3) and 2 (1-5), respectively (Table 2). The magnitude of the difference in the average raw scores between the revised and original UPSIT significantly differed between control and PD participants (*P* = .0466). Scores were not impacted by test order.

Percentile scores were higher with the revised UPSIT compared to the original in both study groups; the median (IQR) percentile difference was 13.5 (1.5-27) among HC, and only 2 (0-6) in the PD cohort (Table 2). The magnitude of the difference in the percentile scores between the revised and original UPSIT significantly differed between HC and PD participants (*P* = .0019).

### Agreement between original and revised UPSIT raw scores

A Bland-Altman plot is used in Figure 1 to visualize the level of agreement between the original and revised versions of the UPSIT. A systematic difference is observed in which scores on the revised UPSIT are generally higher (mean: 2) than on the original UPSIT. The figure also visualizes the finding that, on average, HC participants had higher UPSIT scores than PD participants.

**Figure 1.**
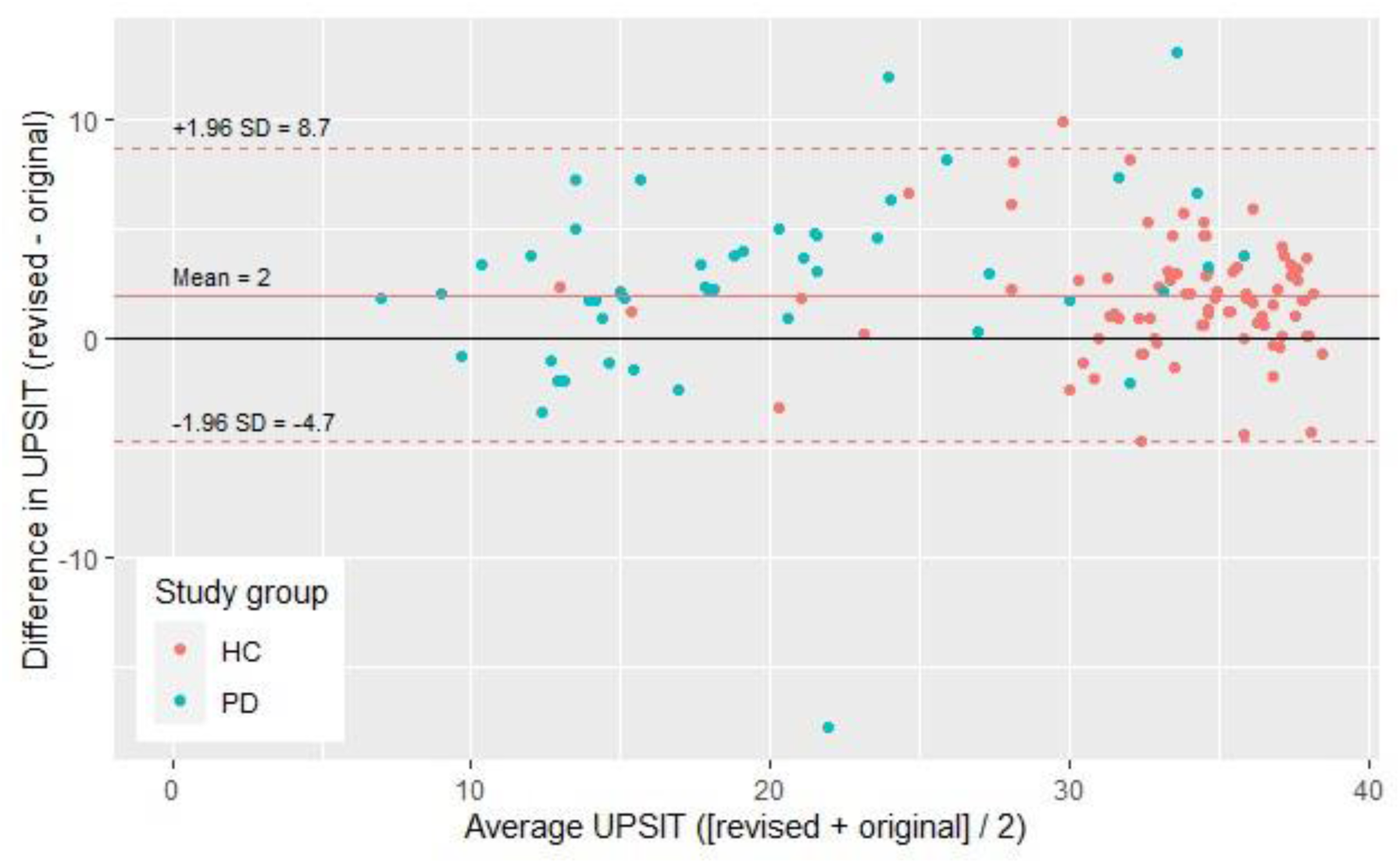
Bland-Altman plot demonstrating the relationship between the average UPSIT score for each participant in the bridging study (x-axis) and the difference in UPSIT raw scores (y-axis). PD participants are plotted in blue; HC participants are plotted in orange.

### Cohort Study to Derive Updated Percentiles: Between-subject comparison of percentile scores Characteristics of the study population

Original UPSIT raw scores among 4,246 females (mean [SD] age: 67.8 [6.1]) and 3,679 males (69.7 [6.7]) aged ≥60 years from the previously published PARS/PPMI combined cohort^9^ were compared to the raw scores of the revised UPSIT among two newly enrolled cohorts. The new cohorts included 16,972 total individuals: 2,485 females (mean [SD] age: 68.0 [5.6]) and 1,908 males (69.6 [6.2]) recruited online (PPMI Online); and 9,269 females (68.0 [5.9]) and 3,310 males (69.9 [6.5]) aged ≥60 who completed the revised UPSIT through ST Direct.

Demographics of this study population are shown in Table 3. The ST Direct population (n=12,579) was much larger than the PPMI Online (n=4,393) population; demographic characteristics were found to be evenly matched in the two populations (i.e., age, distribution by age category, race, and ethnicity). The proportion with a family history of PD differed between the groups as a function of the recruitment methods; 20.0% of the ST Direct population and 47.8% of the PPMI Online population reported a family history of PD.

**Table 3.** Demographics of participants in cohort study used to derive updated percentiles for the revised UPSIT.

| Variable | Females by cohort |  |  | Males by cohort |  |  |
| --- | --- | --- | --- | --- | --- | --- |
|  | Combined<br>(N = 11754) | PPMI Online<br>(N = 2485) | ST Direct<br>(N = 9269) | Combined<br>(N = 5218) | PPMI Online<br>(N = 1908) | ST Direct<br>(N = 3310) |
| **Age, mean (SD)** | 68.0 (5.8) | 68.0 (5.6) | 68.0 (5.9) | 69.8 (6.4) | 69.6 (6.2) | 69.9 (6.5) |
| Median (min, max) | 67.0 (60, 96) | 67.4 (60, 92) | 66.9 (60, 96) | 69.1 (60, 95) | 69.2 (60, 94) | 69.1 (60, 95) |
| **Age category, n (%)** |  |  |  |  |  |  |
| 60-64 | 4266 (36%) | 878 (35%) | 3388 (37%) | 1363 (26%) | 486 (25%) | 877 (26%) |
| 65-69 | 3673 (31%) | 746 (30%) | 2927 (32%) | 1517 (29%) | 560 (29%) | 957 (29%) |
| 70-74 | 2299 (20%) | 538 (22%) | 1761 (19%) | 1265 (24%) | 487 (26%) | 778 (24%) |
| 75-79 | 1072 (9%) | 255 (10%) | 817 (9%) | 718 (14%) | 257 (13%) | 461 (14%) |
| 80+ | 444 (4%) | 68 (3%) | 376 (4%) | 355 (7%) | 118 (6%) | 237 (7%) |
| **Race (White)<sup>a</sup>, n (%)** | 11323 (97%) | 2379 (98%) | 8944 (97%) | 4969 (97%) | 1818 (97%) | 3151 (97%) |
| Missing | 112 | 47 | 65 | 93 | 37 | 56 |
| **Ethnicity (Hispanic), n (%)** | 225 (2%) | 43 (2%) | 182 (2%) | 119 (2%) | 41 (2%) | 78 (2%) |
| Missing | 431 | 19 | 412 | 188 | 14 | 174 |
| **Family history of PD, n (%)** |  |  |  |  |  |  |
| No | 7845 (67%) | 1044 (42%) | 6801 (74%) | 3619 (70%) | 1088 (57%) | 2531 (77%) |
| Yes | 3286 (28%) | 1360 (55%) | 1926 (21%) | 1310 (25%) | 741 (39%) | 569 (17%) |
| Not sure | 532 (5%) | 80 (3%) | 452 (5%) | 247 (5%) | 79 (4%) | 168 (5%) |
| Missing | 91 | 1 | 90 | 42 | 0 | 42 |

<sup>a</sup> Across PPMI Online and ST Direct, 30 individuals self-reported as American Indian/Alaska Native, 142 as Asian, 114 as Black or African American, 7 as Native Hawaiian/Pacific Islander, and 182 as multiracial.
PD = Parkinson's disease; PPMI = Parkinson's Progression Markers Initiative.

The PPMI Online and ST Direct populations were similar in age, race and ethnicity as the PPMI and PARS cohorts previously reported.^9^ In the PARS/PPMI combined cohort, 34% reported a first-degree relative with PD, as compared to 19.8% and 47.8% in the ST Direct and PPMI Online cohorts, respectively. Although not diagnosed with PD, some prodromal features were reported by study participants. Specifically, 12% of female and 10% of male PPMI Online participants self-reported regular (at least weekly) use of laxatives; 23% of female and 18% of male PPMI Online participants endorsed constipation (i.e., <1 bowel movement per day).

Symptoms of REM sleep behavior disorder (RBD) were self-reported by 10% of female and 22% of male participants in the combined PPMI Online/ST Direct cohort, with 2% and 6% of females and males, respectively, having received a diagnosis of RBD.

Additional data for these demographic parameters including the prodromal features by the separate cohorts are provided in the supplementary materials for this article (Table S1).

### Revised UPSIT percentiles

Percentile scores for the revised UPSIT derived from the newly enrolled PPMI Online/ST Direct cohort are presented in Table 4 for females and Table 5 for males, broken down into five age categories (i.e., 60-64, 65-69, 70-74, 75-79, and ≥80 years).

**Table 4.** Age-specific UPSIT percentile values among females.

| UPSIT Raw Score | Percentile by Age Category |  |  |  |  |
| --- | --- | --- | --- | --- | --- |
|  | 60-64 y | 65-69 y | 70-74 y | 75-79 y | ≥80 y |
| 40 | 98 | 98.5 | 99 | 99.5 | >99.5 |
| 39 | 88.5 | 91 | 93.5 | 96 | 98 |
| 38 | 73.5 | 78.5 | 83 | 89 | 93.5 |
| 37 | 57 | 64 | 69.5 | 79 | 86.5 |
| 36 | 42 | 50 | 56 | 67.5 | 77.5 |
| 35 | 31 | 38.5 | 45 | 56 | 67.5 |
| 34 | 23.5 | 30 | 36.5 | 46 | 58.5 |
| 33 | 18 | 24 | 29.5 | 38 | 50.5 |
| 32 | 14 | 19.5 | 24.5 | 32.5 | 43.5 |
| 31 | 11.5 | 16.5 | 21 | 29 | 38.5 |
| 30 | 10 | 14 | 18 | 25.5 | 34 |
| 29 | 9 | 12 | 15.5 | 22 | 29.5 |
| 28 | 8 | 11 | 13.5 | 19.5 | 26.5 |
| 27 | 7 | 10 | 12 | 18 | 23 |
| 26 | 6 | 9 | 11 | 16.5 | 20 |
| 25 | 6 | 8 | 10 | 14.5 | 18 |
| 24 | 5 | 7 | 9 | 13 | 16.5 |
| 23 | 5 | 6 | 9 | 12 | 15 |
| 22 | 4 | 5 | 8 | 12 | 13.5 |
| 21 | 4 | 5 | 7 | 10.5 | 12 |
| 20 | 3 | 4 | 6 | 8.5 | 11 |
| 19 | 3 | 4 | 5 | 7 | 10 |
| 18 | 3 | 3 | 4 | 6 | 10 |
| 17 | 2 | 3 | 4 | 6 | 9 |
| 16 | 2 | 2 | 3 | 4.5 | 8 |
| 15 | 2 | 2 | 3 | 3 | 7 |

| Percentile by Age Category |  |  |  |  |  |
| --- | --- | --- | --- | --- | --- |
| UPSIT Raw Score | 60-64 y | 65-69 y | 70-74 y | 75-79 y | ≥80 y |
| 14 | 2 | 2 | 2 | 3 | 6 |
| 13 | 1 | 1 | 2 | 2 | 5 |
| 12 | 1 | 1 | 2 | 2 | 3.5 |
| 11 | 1 | 1 | 1 | 1 | 2 |
| 10 | 1 | 1 | 1 | 1 | 1 |
| <10 | 1 | 1 | 1 | 1 | 1 |
| N | 4266 | 3673 | 2299 | 1072 | 444 |
| UPSIT Score, Mean (SD) | 35.2 (5.0) | 34.3 (5.5) | 33.5 (6.1) | 32.1 (6.7) | 30.5 (7.3) |
| UPSIT Score, Median (Min, Max) | 37.0 (3, 40) | 36.0 (7, 40) | 36.0 (4, 40) | 34.0 (8, 40) | 33.0 (4, 40) |

<sup>a</sup> If a raw UPSIT score corresponded to multiple percentile values, the median percentile value was selected; as a result, some percentile values are non-integers.
UPSIT = University of Pennsylvania Smell Identification Test.

**Table 5.** Age-specific UPSIT percentile values among males.

| UPSIT Score | Percentile by Age Category |  |  |  |  |
| --- | --- | --- | --- | --- | --- |
|  | 60-64 y | 65-69 y | 70-74 y | 75-79 y | ≥80 y |
| 40 | 98.5 | 99.5 | 99.5 | >99.5 | >99.5 |
| 39 | 92 | 95.5 | 97 | 98 | 98.5 |
| 38 | 81 | 88 | 92 | 94.5 | 95.5 |
| 37 | 68 | 77.5 | 84 | 89 | 91.5 |
| 36 | 55.5 | 65 | 74 | 80.5 | 86 |
| 35 | 45 | 53.5 | 63.5 | 70.5 | 79.5 |
| 34 | 36 | 44.5 | 54 | 63 | 73.5 |
| 33 | 29 | 37 | 45.5 | 56 | 68 |
| 32 | 25 | 31 | 39 | 48.5 | 63.5 |
| 31 | 22.5 | 26.5 | 34.5 | 43 | 59 |
| 30 | 20 | 23 | 31 | 38 | 54.5 |
| 29 | 18 | 20.5 | 28 | 34 | 51 |
| 28 | 16.5 | 19 | 25.5 | 31 | 47 |
| 27 | 15 | 17.5 | 23.5 | 28.5 | 43 |
| 26 | 13.5 | 16 | 21.5 | 25.5 | 40 |
| 25 | 12 | 15 | 19.5 | 22.5 | 37.5 |
| 24 | 10.5 | 14 | 17.5 | 21 | 34.5 |
| 23 | 9 | 13 | 15.5 | 19.5 | 30 |
| 22 | 8 | 11.5 | 14 | 17.5 | 26 |
| 21 | 8 | 11 | 13 | 16 | 24 |
| 20 | 7 | 9.5 | 11.5 | 15 | 22.5 |
| 19 | 6 | 8 | 10 | 15 | 20.5 |
| 18 | 5 | 7 | 9 | 14 | 18.5 |
| 17 | 4 | 6 | 8 | 12.5 | 16 |
| 16 | 4 | 5 | 7 | 10.5 | 14 |
| 15 | 4 | 4 | 6 | 8.5 | 12.5 |

| Percentile by Age Category |  |  |  |  |  |
| --- | --- | --- | --- | --- | --- |
| UPSIT Score | 60-64 y | 65-69 y | 70-74 y | 75-79 y | ≥80 y |
| 14 | 3 | 3 | 5 | 6.5 | 10.5 |
| 13 | 2 | 2 | 4 | 5 | 8 |
| 12 | 2 | 2 | 3 | 4 | 5.5 |
| 11 | 1 | 1 | 2 | 3 | 3.5 |
| 10 | 1 | 1 | 1 | 1.5 | 1.5 |
| <10 | 1 | 1 | 1 | 1 | 1 |
| N | 1363 | 1517 | 1265 | 718 | 355 |
| UPSIT Score, Mean (SD) | 33.3 (6.5) | 32.2 (6.9) | 30.9 (7.4) | 29.6 (7.8) | 27.1 (8.4) |
| UPSIT Score, Median (Min, Max) | 36.0 (2, 40) | 35.0 (7, 40) | 34.0 (4, 40) | 32.0 (8, 40) | 29.0 (2, 40) |

<sup>a</sup> If a raw UPSIT score corresponded to multiple percentile values, the median percentile value was selected; as a result, some percentile values are non-integers.
UPSIT = University of Pennsylvania Smell Identification Test.

A comparison of the percentiles for the original (PARS/PPMI cohorts) and revised (PPMI Online/ST Direct cohorts) UPSIT is provided in Table 6. Across all age and sex categories, UPSIT mean raw scores were higher with the revised compared to the original UPSIT, but the magnitude of the difference varied by age and sex. The smallest mean differences (range 0.6-1.2) were observed among the three oldest (70-74, 75-79, ≥80) male categories and the oldest (≥80) female category. The largest mean differences (range: 1.9-2.1) were evident among the three youngest (60-64, 65-69, 70-74) female categories and the youngest (60-64) male category. When comparing differences in the raw scores that defined key percentile cutoff values (i.e., ≤25^th^, ≤20^th^, ≤15^th^, and ≤10^th^ percentile) for the original and revised UPSIT percentiles, the greatest differences were observed in females; several of the male subgroups had no change or a worsening (i.e., a lower raw score defining the cutoff) for a given percentile with the revised UPSIT.

**Table 6.**
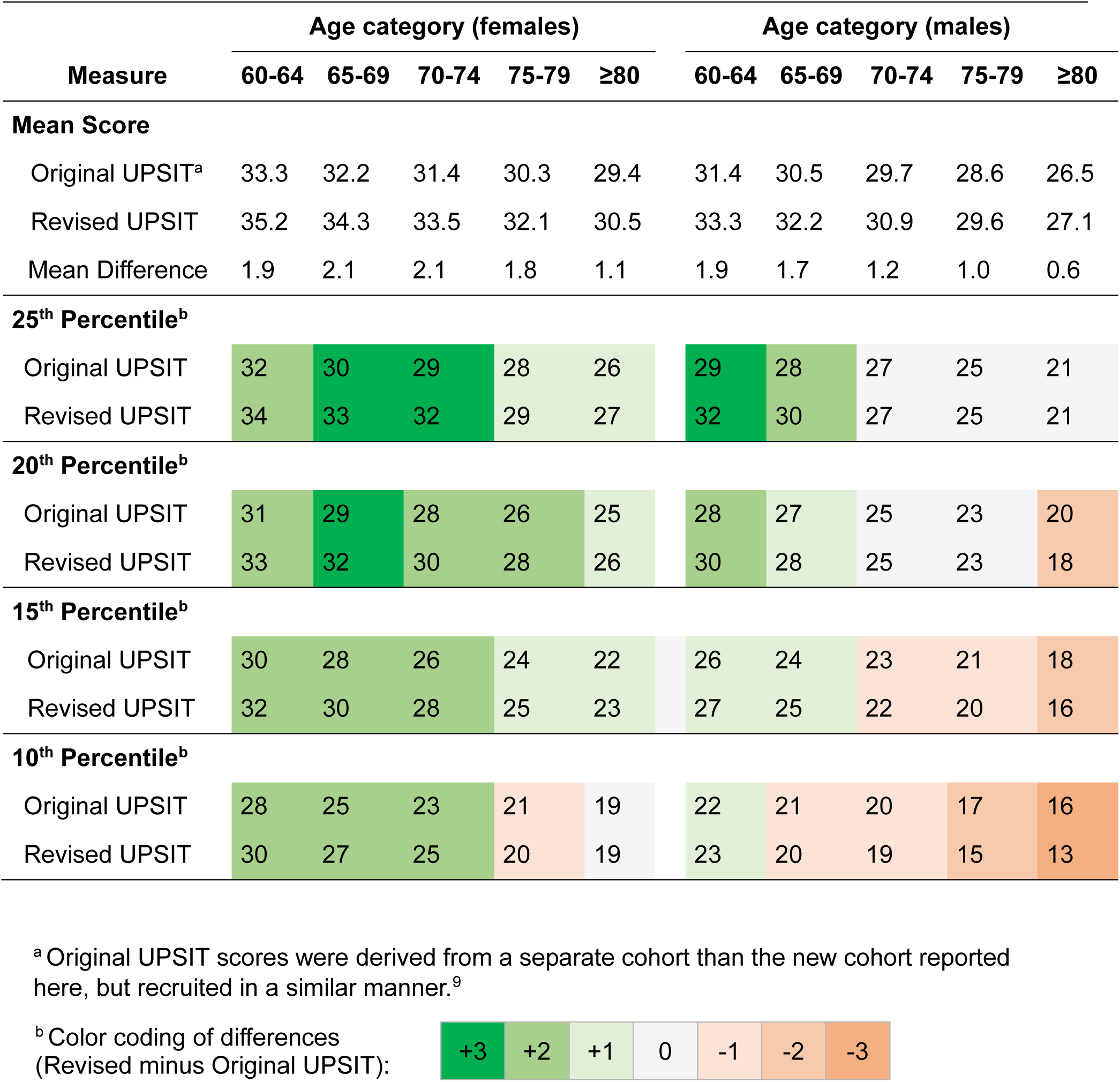
Comparison of original and revised UPSIT raw scores and percentile values by sex and age (cohort study)

## Discussion

As assessment tools evolve over time, so too must our understanding of the data provided by these tools. The measurement of olfactory function is an important prognostic marker of neurodegeneration, and thus it is important for neurologists or any researcher who investigates olfaction to ensure accurate interpretation of such data. The UPSIT has been a mainstay in the evaluation of olfaction, and, after nearly 4 decades of use, has been updated to remain current.

This investigation compares participant performance on the original UPSIT from 1984 to the revised version from 2020 and provides three main insights. First, better performance (i.e., higher scores indicating greater olfactory function) was observed with the revised test compared to the original test. This is an expected finding, consistent with the intention of the revision to modernize the test through updated odorants and distractor responses. Second, this report provides age- and sex-specific percentile look-up tables for the revised UPSIT, which complement the tables previously published for the original version of the test.^9^ Third, of particular relevance to investigators of neurodegeneration, the deficit of olfactory function in PD participants compared to HC participants persists with the revised version of the UPSIT as it has been consistently reported with the original version.

While the main finding of this study is that the revised UPSIT yields slightly higher raw scores, upon closer examination of differences between the original and revised versions of the UPSIT, some nuanced observations can be made. The magnitude of the differences varied by age and sex, with smaller mean differences observed with increasing age, and the larger mean differences observed in females compared to males. The within-subject design of the bridging study allows for the most direct comparison of test performance with the two versions of the UPSIT. On average there is approximately a 2-point improvement in raw scores on the revised UPSIT compared to the original (controls mean difference: 1.7, PD mean difference: 2.5).

However, again this finding is nuanced; what appear to be rather consistent differences in mean scores translates into a greater observed difference in percentile scores (controls percentile mean difference: 14.7, PD percentile mean difference 7.9).

Taken together, these observations highlight the importance of referring to the revised percentile tables provided herein when scoring the revised UPSIT in clinical or research settings. Our findings suggest that it would not be accurate to refer to the percentile look-up tables previously reported for the original version when using the revised test. While raw scores on the revised UPSIT are generally 2-3 points higher than on the original, there are subtle differences based on age and sex that make the granular data provided by our large cohort the best reference for deriving percentiles for the revised UPSIT in cohorts such as ours. The results of the within-subjects bridging study (i.e., median +2-point difference between original and revised UPSIT scores) are consistent with a comparison of the original and revised UPSIT using the large cohorts to generate the percentiles (see comparisons in Table 6), leveraging our original work on percentile score published in 2023,^9^ giving further support for our estimates for age- and sex-specific percentiles.

As with the previous report, this work is strengthened by the large cohort size. Data from a total of 9,396 individuals were analyzed in the prior study of the original UPSIT^9^; an even larger total cohort of 16,972 was analyzed here from PPMI Online and ST Direct for the purposes of generating percentile scores for the revised UPSIT. Because these percentile look-up tables are age- and sex-specific, it is important to have a very robust sample size so that each individual cell (age group by sex) is of sufficient size to reliably estimate corresponding percentiles.

Although we do have a direct comparison of original and revised UPSIT scores within the large cohort, the recruitment methods used in this study and our prior work^9^ are similar enough to expect that the results would not be dramatically different. The concordant results in the smaller within-subject component of this report support this expectation.

Despite the robust sample size, this study is limited by a lack of racial and ethnic diversity; the cohort study participants self-reported as 97% White and 98% non-Hispanic. It was anticipated that the large sample size combined with the broad recruitment strategies (particularly for ST Direct) would provide a more diverse cohort as compared to previous efforts. However, this was not the case. Although the sample may be inadequate to draw conclusions based on race/ethnicity, the reader is referred to the supplementary materials for this article to review the demographic features and UPSIT raw and percentile scores by self-reported race (Table S2) and ethnicity (Table S3) for this study. The bridging study did introduce perhaps a somewhat more diverse cohort based on purposeful sampling, and 17% (n=9) of the PD participants were self-reported persons of color. Nevertheless, it is clear that the findings reported here are most generalizable to White, non-Hispanic populations. Known differences in odorant recognition and prevalence of olfactory dysfunction by race and ethnicity have been documented,^13, 14, 15, 16, 17, 18, 12–17^ thereby making it a priority to investigate olfactory function in much more diverse cohorts.

Several limitations pertain to the within-subjects design employed in the bridging study. There is an uneven distribution of participants by site and by disease status. Specifically, 100% of the PD participants were recruited at Penn and a large majority of the HC participants (89%) were recruited at IU. Thus, differences in UPSIT performance attributed to disease status (HC vs. PD) could be driven by unrecognized site differences. However, we did not observe substantial differences between the results for normosmic subjects recruited at Penn compared to those recruited through IU. In addition, in the IU participants, all completed the original test first and the revised test second. However, average performance improved on the revised test, suggesting that a change in olfactory function during the inter-test interval was not likely to account for the observed effect.

In summary, our results provide a reliable and definitive basis for converting raw scores on the revised UPSIT into percentiles that are appropriate for screening older populations for neurodegenerative disorders such as PD, and more broadly, neuronal synuclein disease.^19^ The case has previously been made for the utility of percentiles over threshold values for the assessment of olfactory function.^9^ Therefore, it is hoped that this report, and specifically the inclusion of percentile look-up tables for the revised UPSIT (2020), helps support ongoing and future investigations of the role of olfactory function to aid interpretation of findings, particularly among persons with neurodegenerative diseases. In PD, this work is critical to the mission of PPMI to identify markers of disease progression and, in particular, enable a deeper investigation for the important role for olfactory function in that pursuit.

## Data Availability

This analysis used data openly available from PPMI; data used in the preparation of this manuscript were obtained on July 3, 2023 from the Parkinson's Progression Markers Initiative (PPMI) database (www.ppmi-info.org/access-data-specimens/download-data), RRID: SCR 006431. For up-to-date information on the study, visit www.ppmi-info.org. This analysis was conducted by the PPMI Statistics Core and used actual dates of activity for participants, a restricted data element not available to public users of PPMI data. Statistical analysis codes used to perform the analyses in this article are shared on Zenodo [10.5281/zenodo.11644053].

## Acknowledgements

The authors sincerely appreciate the numerous and valuable contributions of the large number of study participants in PPMI and ST Direct, as well as the study teams.

## Collaborators

University of Pennsylvania (alphabetical): Justin James, Crystalle Hutchins, Suzanne Reichwein, and Ian Walker.

Indiana University (alphabetical): Isaac Harms, Christopher Hobbick, Joseph McClaren

## Executive Steering Committee

Kenneth Marek, MD^1^ (Principal Investigator); Caroline Tanner, MD, PhD^9^; Tanya Simuni, MD^3^; Andrew Siderowf, MD, MSCE^12^; Douglas Galasko, MD^27^; Lana Chahine, MD^39^; Christopher Coffey, PhD^4^; Kalpana Merchant, PhD^59^; Kathleen Poston, MD^38^; Roseanne Dobkin, PhD^41^; Tatiana Foroud, PhD^15^; Brit Mollenhauer, MD^8^; Dan Weintraub, MD^12^; Ethan Brown, MD^9^; Karl Kieburtz, MD, MPH^23^; Mark Frasier, PhD^6^; Todd Sherer, PhD^6^; Sohini Chowdhury, MA^6^; Roy Alcalay, MD^35^ and Aleksandar Videnovic, MD^45^

## Steering Committee

Duygu Tosun-Turgut, PhD^9^; Werner Poewe, MD^7^; Susan Bressman, MD^14^; Jan Hammer^15^; Raymond James, RN^22^; Ekemini Riley, PhD^40^; John Seibyl, MD^1^; Leslie Shaw, PhD^12^; David Standaert, MD, PhD^18^; Sneha Mantri, MD, MS^60^; Nabila Dahodwala, MD^12^; Michael Schwarzschild^45^; Connie Marras^43^; Hubert Fernandez, MD^25^; Ira Shoulson, MD^23^; Helen Rowbotham^2^; Paola Casalin^11^ and Claudia Trenkwalder, MD^8^

## Michael J. Fox Foundation (Sponsor)

Todd Sherer, PhD; Sohini Chowdhury, MA; Mark Frasier, PhD; Jamie Eberling, PhD; Katie Kopil, PhD; Alyssa O’Grady; Maggie McGuire Kuhl; Leslie Kirsch, EdD; and Tawny Willson, MBS

## Study Cores, Committees and Related Studies

*SPPMI Online:* Carlie Tanner, MD PhD^9^; Ethan Brown, MD^9^; Lana Chahine, MD^39^; Roseanne Dobkin, PhD^41^; Monica Korell, MPH^9^

*Screening Core:* Tatiana Foroud, PhD^15^; Laura Heathers, BA^15^; Christopher Hobbick, BSCE^15^; Gena Antonopoulos, BSN^15^

*Statistics Core*: Christopher Coffey, PhD^4^; Chelsea Caspell-Garcia, MS^4^; Michael Brumm, MS^4^

## Site Investigators

Charles Adler, PhD^49^; Roy Alcalay, MD^35^; Amy Amara, PhD^50^; Paolo Barone, PhD^30^; Bastiaan Bloem, PhD^58^; Susan Bressman, MD^14^; Kathrin Brockmann, MD^26^; Norbert Brüggemann, MD^57^; Lana Chahine, MD^39^; Kelvin Chou, MD^42^; Nabila Dahodwala, MD^12^; Alberto Espay, MD^32^; Stewart Factor, DO^16^; Hubert Fernandez, MD^25^; Michelle Fullard, MD^50^; Douglas Galasko, MD^27^; Robert Hauser, MD^19^; Penelope Hogarth, MD^17^; Shu-Ching Hu, PhD^21^; Michele Hu, PhD^56^; Stuart Isaacson, MD^31^; Christine Klein, MD^57^; Rejko Krueger, MD^2^; Mark Lew, MD^47^; Zoltan Mari, MD^54^; Connie Marras, PhD^43^; Maria Jose Martí, PhD^33^; Nikolaus McFarland, PhD^52^; Tiago Mestre, PhD^44^; Brit Mollenhauer, MD^8^; Emile Moukheiber, MD^28^; Alastair Noyce, PhD^61^; Wolfgang Oertel, PhD^62^; Njideka Okubadejo, MD^63^; Sarah O’Shea, MD^37^; Rajesh Pahwa, MD^46^; Nicola Pavese, PhD^55^; Werner Poewe, MD^7^; Ron Postuma, MD^53^; Giulietta Riboldi, MD^51^; Lauren Ruffrage, MS^18^; Javier Ruiz Martinez, PhD^34^; David Russell, PhD^1^; Marie H Saint-Hilaire, MD^22^; Neil Santos, BS^49^; Wesley Schlett^45^; Ruth Schneider, MD^23^; Holly Shill, MD^48^; David Shprecher, DO^24^; Tanya Simuni, MD^3^; David Standaert, PhD^18^; Leonidas Stefanis, PhD^36^; Yen Tai, PhD^29^; Caroline Tanner, PhD^9^; Arjun Tarakad, MD^20^; Eduardo Tolosa PhD^33^ and Aleksandar Videnovic, MD^45^

## Coordinators

Susan Ainscough, BA^30^; Courtney Blair, MA^18^; Erica Botting^19^; Isabella Chung, BS^54^; Kelly Clark^24^; Ioana Croitoru^34^; Kelly DeLano, MS^32^; Iris Egner, PhD^7^; Fahrial Esha, BS^51^; May Eshel^35^; Frank Ferrari, BS^42^; Victoria Kate Foster^55^; Alicia Garrido, MD^33^; Madita Grümmer^57^; Bethzaida Herrera^48^; Ella Hilt^26^; Chloe Huntzinger, BA^50^; Raymond James, BS^22^; Farah Kausar, PhD^9^; Christos Koros, MD, PhD^36^; Yara Krasowski^58^; Dustin Le, BS^17^; Ying Liu, MD^50^; Taina M. Marques, PhD^2^; Helen Mejia Santana, MA^37^; Sherri Mosovsky, MPH^39^; Jennifer Mule, BS^25^; Philip Ng, BS^43^; Lauren O’Brien^46^; Abiola Ogunleye, PGDip^29^; Oluwadamilola Ojo, MD^63^; Obi Onyinanya, BS^28^; Lisbeth Pennente, BA^31^; Romina Perrotti^53^; Michael Pileggi, MS^53^; Ashwini Ramachandran, MSc^12^; Deborah Raymond, MS^14^; Jamil Razzaque, MS^56^; Shawna Reddie, BA^44^; Kori Ribb, BSN^28^; Kyle Rizer, BA^52^; Janelle Rodriguez, BS^27^; Stephanie Roman, HS^1^; Clarissa Sanchez, MPH^20^; Cristina Simonet, PhD^29^; Anisha Singh, BS^23^; Elisabeth Sittig^62^; Barbara Sommerfeld MSN^16^; Angela Stovall, BS^42^; Bobbie Stubbeman, BS^32^; Alejandra Valenzuela, BS^47^; Catherine Wandell, BS^21^; Diana Willeke^8^; Karen Williams, BA^3^ and Dilinuer Wubuli, MB^43^

1 Institute for Neurodegenerative Disorders, New Haven, CT

2 University of Luxembourg, Luxembourg

3 Northwestern University, Chicago, IL

4 University of Iowa, Iowa City, IA

5 VectivBio AG

6 The Michael J. Fox Foundation for Parkinson’s Research, New York, NY

7 Innsbruck Medical University, Innsbruck, Austria

8 Paracelsus-Elena Klinik, Kassel, Germany

9 University of California, San Francisco, CA

10 Laboratory of Neuroimaging (LONI), University of Southern California

11 BioRep, Milan, Italy

12 University of Pennsylvania, Philadelphia, PA

13 National Institute on Aging, NIH, Bethesda, MD

14 Mount Sinai Beth Israel, New York, NY

15 Indiana University, Indianapolis, IN

16 Emory University of Medicine, Atlanta, GA

17 Oregon Health and Science University, Portland, OR

18 University of Alabama at Birmingham, Birmingham, AL

19 University of South Florida, Tampa, FL

20 Baylor College of Medicine, Houston, TX

21 University of Washington, Seattle, WA

22 Boston University, Boston, MA

23 University of Rochester, Rochester, NY

24 Banner Research Institute, Sun City, AZ

25 Cleveland Clinic, Cleveland, OH

26 University of Tübingen, Tübingen, Germany

27 University of California, San Diego, CA

28 Johns Hopkins University, Baltimore, MD

29 Imperial College of London, London, UK

30 University of Salerno, Salerno, Italy

31 Parkinson’s Disease and Movement Disorders Center, Boca Raton, FL

32 University of Cincinnati, Cincinnati, OH

33 Hospital Clinic of Barcelona, Barcelona, Spain

34 Hospital Universitario Donostia, San Sebastian, Spain

35 Tel Aviv Sourasky Medical Center, Tel Aviv, Israel

36 National and Kapodistrian University of Athens, Athens, Greece

37 Columbia University Irving Medical Center, New York, NY

38 Stanford University, Stanford, CA

39 University of Pittsburgh, Pittsburgh, PA

40 Center for Strategy Philanthropy at Milken Institute, Washington D.C.

41 Rutgers University, Robert Wood Johnson Medical School, New Brunswick, New Jersey

42 University of Michigan, Ann Arbor, MI

43 Toronto Western Hospital, Toronto, Canada

44 The Ottawa Hospital, Ottawa, Canada

45 Massachusetts General Hospital, Boston, MA

46 University of Kansas Medical Center, Kansas City, KS

47 University of Southern California, Los Angeles, CA

48 Barrow Neurological Institute, Phoenix, AZ

49 Mayo Clinic Arizona, Scottsdale, AZ

50 University of Colorado, Aurora, CO

51 NYU Langone Medical Center, New York, NY

52 University of Florida, Gainesville, FL

53 Montreal Neurological Institute and Hospital/McGill, Montreal, QC, Canada

54 Cleveland Clinic-Las Vegas Lou Ruvo Center for Brain Health, Las Vegas, NV

55 Clinical Ageing Research Unit, Newcastle, UK

56 John Radcliffe Hospital Oxford and Oxford University, Oxford, UK

57 Universität Lübeck, Luebeck, Germany

58 Radboud University, Nijmegen, Netherlands 59 TransThera Consulting

60 Duke University, Durham, NC

61 Wolfson Institute of Population Health, Queen Mary University of London, UK

62 Philipps-University Marburg, Germany

63 University of Lagos, Nigeria

## Supplementary Materials

Supplementary Table S1. Comparison of demographics and prodromal features of cohort study participants

Supplementary Table S2. UPSIT percentile by self-reported race

Supplementary Table S3. UPSIT percentile by self-reported ethnicity

